# COVID-19 vaccine acceptance among healthcare workers in India: Results from a cross-sectional survey

**DOI:** 10.1101/2021.08.10.21261868

**Authors:** Kayur Mehta, Baldeep K. Dhaliwal, Sanjay Zodpey, Stacie Loisate, Preetika Banerjee, Madhu Gupta, Anita Shet

**Author notes:** **Address correspondence to:** Dr. Anita Shet, MD, PhD., Director, Maternal and Child Health Center India, International Vaccine Access Center, 415 N Washington Street 5th Floor, Baltimore, MD 21231. Authors contributed equally to this work (Kayur Mehta, Baldeep K. Dhaliwal).

## Abstract

**Background:** Remarkable scientific progress has enabled expeditious development of effective vaccines against COVID-19. While healthcare workers (HCWs) have been at the frontlines for the pandemic response, vaccine acceptance amongst them needs further study.

**Methods:** A web-based survey to assess vaccine acceptance and preparedness in India was disseminated to HCWs working in various settings between January and February 2021, shortly after the launch of India’s vaccination campaign. Descriptive statistics were used to examine respondent demographics and Likert scale responses. Binomial logistic regression analyses were used to identify factors associated with vaccine acceptance.

**Results:** The survey yielded 624 respondents from 25 states and five union territories in India; 53.5% were male, and median age was 37 years (IQR 32-46). Amongst all respondents, 84.1% (525/624) supported COVID-19 vaccines, and 63.2% (141/223) of those unvaccinated at the time of survey administration were willing to accept a vaccine. Reliability on government sources, healthcare providers or scientific journal articles for COVID-19 related information was reported by 66.8%, while confidence in social media for this information was reported by only 4.5%. Factors independently associated with vaccine acceptance included advancing age (aOR 3.50 [95% CI, 1.04-11.76] for those above 45 years), evidence of vaccine effectiveness and safety (aOR 3.78 [95% CI 1.15-12.38]), and provision of free/no-cost vaccine (aOR 2.63 [95% CI, 1.06-6.50]). Most respondents (80%) were confident about their hospital being equipped to efficiently rollout COVID-19 vaccines to the general population.

**Conclusions:** Overall attitudes towards COVID-19 vaccination and preparedness were positive among HCWs in India, although acceptance was lower among healthier and younger HCWs. Data availability on vaccine safety and effectiveness, and cost considerations were important for acceptance. Targeted interventions are needed to improve vaccine acceptance amongst HCWs, since they are critical in promoting vaccine acceptance amongst the general population.

## Background

After first emerging in late 2019 (1), the SARS-CoV-2 virus which causes COVID-19 has spread across the world and achieved pandemic status in March 2020. In the absence of pharmaceutical interventions, population-wide lockdowns and social distancing measures were enforced to slow the spread of the virus and reduce deaths (2). However, rapidly developing an efficacious vaccine was crucial to the control and mitigation of the pandemic. Unprecedented scientific endeavors saw the development of vaccines against SARS-CoV-2 at extraordinary speed, with clinical trials beginning within two months of the discovery of the SARS-CoV-2 genetic sequence (3). By early January 2021, India’s drug regulator provided emergency use authorization to two vaccine candidates, one manufactured by Serum Institute of India under license from Astra Zeneca, and the other, an inactivated SARS-CoV-2 vaccine manufactured by Bharat Biotech in collaboration with Indian Council of Medical Research (4). Due to their enhanced risk of exposure and infection, healthcare and frontline workers were prioritized for vaccination.

To ensure the success of any vaccination campaign, understanding facilitators and barriers to uptake of the vaccine is critical to address. Following the launch of the COVID-19 vaccination campaign in India, vaccine refusal among healthcare and frontline workers, despite their prioritization, was contributing to low uptake; shortly after the vaccine was authorized, only 56% of eligible HCWs had stepped forward to accept a vaccine (5). Studies from across the world have shown in the context of a COVID-19 vaccine, vaccine hesitancy amongst healthcare workers (HCWs) ranged from 27.7-78.1% (6). Safety concerns, potential adverse events, and speed of vaccine development have been identified as major concerns (6).

HCWs are important sources of information for vaccines, and effective communication from HCWs with patients has been shown to improve adherence to vaccination recommendations (7, 8). Studies have shown that HCWs who are vaccinated are more likely to recommend vaccines to friends, family, and their patients (9, 10). Data regarding COVID-19 vaccine acceptance amongst HCWs in India are scarce. This study aimed to evaluate barriers to acceptance and preparedness to advocate for vaccine uptake among healthcare and frontline workers in India.

## Methods

### Survey questionnaire and design

A survey to assess perceptions of HCWs towards COVID-19 vaccination was developed by researchers at the Johns Hopkins Maternal and Child Health Center in India and the Public Health Foundation of India, using vaccine hesitancy questionnaire development guidelines from the WHO (11), and Kaiser Family Foundation (12). The survey consisted of 34 questions to document demographic information, knowledge and attitudes about COVID-19 and COVID-19 vaccines, perceptions of risk in their work setting, intent to accept a COVID-19 vaccine, and readiness among healthcare facilities to scale up vaccine delivery. Participants could skip any questions they did not wish to answer.

### Participants and survey dissemination

The survey was administered online using the software platform Qualtrics (Qualtrics, Provo, UT) between 24 January 2021 and 28 February 2021. HCWs working in various settings in India were invited to participate after distributing the survey via professional networks such as the Public Health Foundation of India and the Indian Association of Preventive and Social Medicine. The survey was further distributed among administrative leaders across hospitals affiliated with medical colleges and among employees of other major hospital systems in India. Dissemination of the survey was done via email, text messages and social media platforms such as WhatsApp. A snowball sampling method was utilized to increase the reach of the survey. HCWs were classified on the basis of whether or not they worked in a health-facility setting, and those working in health-facility settings were further classified as patient-facing and non-patient facing. Nurses, medical doctors, clinic workers and hospital paramedical workers were considered as patient-facing; non-patient facing HCWs were reception clerks, housekeeping staff, laboratory personnel and other non-healthcare frontline workers. HCWs working as public health professionals outside of health-facility settings were classified as working in non-health-facility settings.

### Statistical Analysis

Descriptive statistics, expressed as frequencies and proportions, were used to examine respondent demographics and Likert scale responses. Proportions were calculated using denominators reflective of the number of respondents for the specific question. Multiple imputation technique was used to correct for missing data in the Likert scale responses. Chi square tests were used to compare the characteristics of respondents who were unwilling to accept a COVID-19 vaccine with those who were willing to, amongst those respondents who had not received a COVID-19 vaccine at the time of the survey. Binomial logistic regression analyses were used to determine factors associated with vaccine acceptance amongst those respondents who reported that they had not yet received any COVID-19 vaccines. Specifically, models were developed to identify characteristics associated with responses of “yes” versus “no” or “I don’t know” (as a composite variable) to whether or not they would be willing to accept a COVID-19 vaccine, thus suggestive of vaccine acceptance. All analyses were conducted using the statistical package R (R Foundation for Statistical Computing, Vienna, Austria. http://www.R-project.org/).

### Ethical considerations

The study was reviewed by the Institutional Ethics Committee at the Public Health Foundation of India and by the Johns Hopkins Bloomberg School of Public Health (JHSPH) Institutional Review Board (IRB) and determined to be exempt from human subjects research oversight. Consent to participate in the survey was provided in the introductory portion of the online survey.

## Results

The online survey was completed by 632 respondents. Amongst these respondents, eight did not respond to the question on intent to be vaccinated which was the primary focus of this survey; following their exclusion, we had a total of 624 respondents, which is the number included in subsequent analyses in this report. These respondents were distributed across 25 states and five union territories in India; 53.5% were male, and the median age was 37 years (IQR 32-46). Amongst the cumulative 624 respondents, 21.5% already had a confirmed COVID-19 diagnosis in the past, and 24.7% reported having underlying medical co-morbidities which would place them at higher risk for severe COVID-19 disease (Table 1).

**Table 1.**
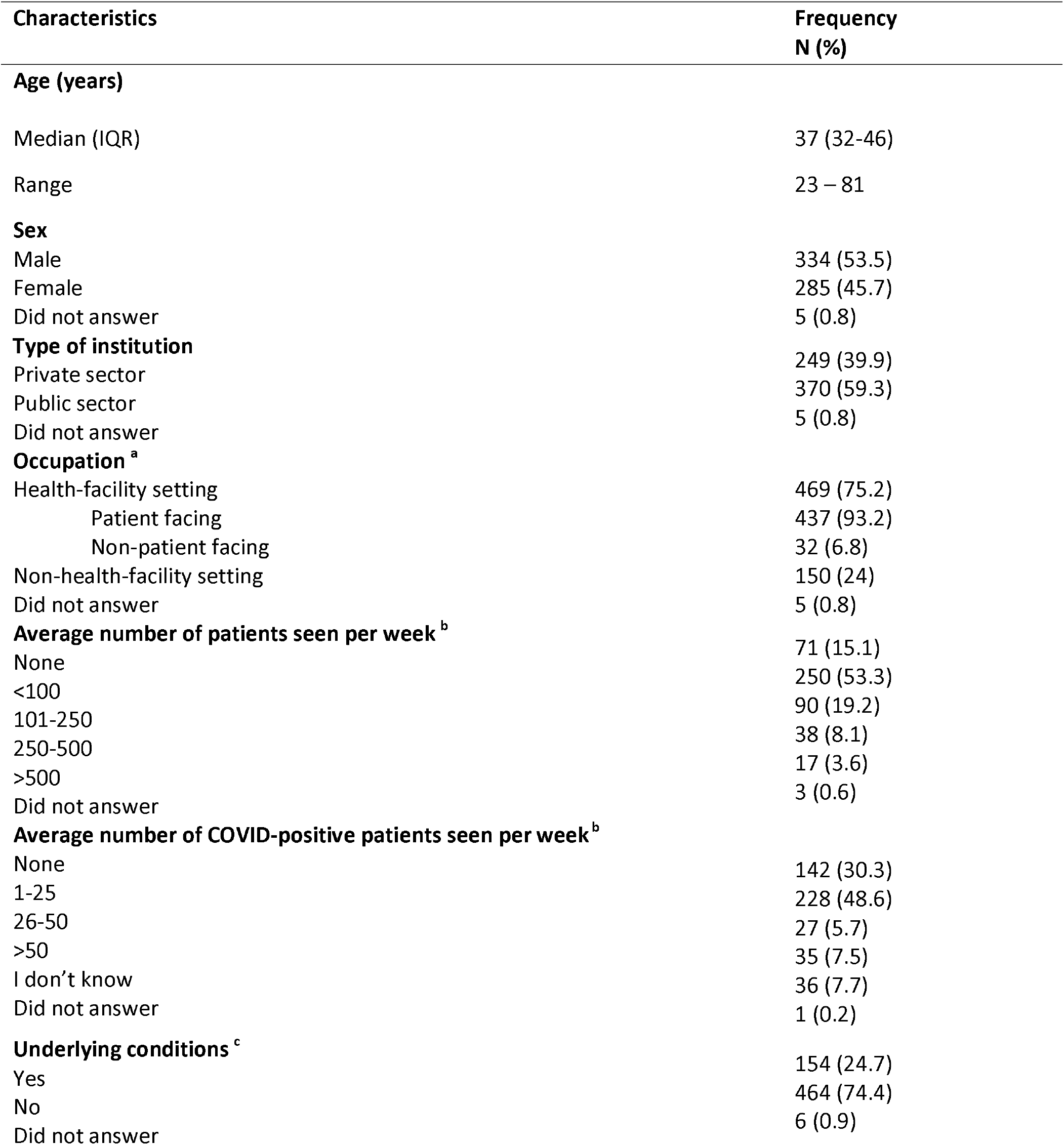
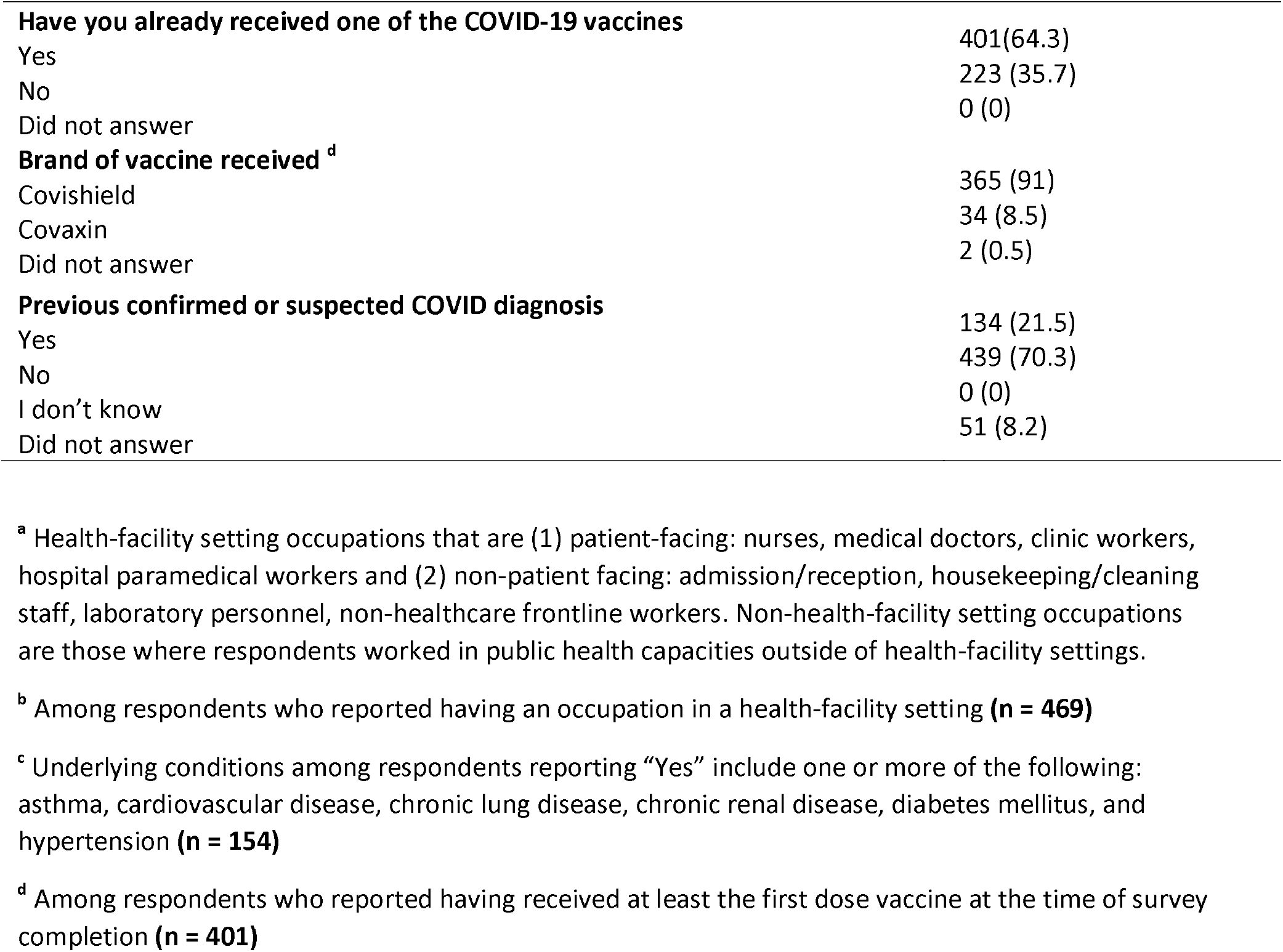
Respondent demographics (n = 624)

| Characteristics | Frequency<br>N (%) |
| --- | --- |
| **Age (years)** |  |
| Median (IQR) | 37 (32-46) |
| Range | 23 – 81 |
| **Sex** |  |
| Male | 334 (53.5) |
| Female | 285 (45.7) |
| Did not answer | 5 (0.8) |
| **Type of institution** |  |
| Private sector | 249 (39.9) |
| Public sector | 370 (59.3) |
| Did not answer | 5 (0.8) |
| **Occupation <sup>a</sup>** |  |
| Health-facility setting | 469 (75.2) |
| Patient facing | 437 (93.2) |
| Non-patient facing | 32 (6.8) |
| Non-health-facility setting | 150 (24) |
| Did not answer | 5 (0.8) |
| **Average number of patients seen per week <sup>b</sup>** |  |
| None | 71 (15.1) |
| <100 | 250 (53.3) |
| 101-250 | 90 (19.2) |
| 250-500 | 38 (8.1) |
| >500 | 17 (3.6) |
| Did not answer | 3 (0.6) |
| **Average number of COVID-positive patients seen per week <sup>b</sup>** |  |
| None | 142 (30.3) |
| 1-25 | 228 (48.6) |
| 26-50 | 27 (5.7) |
| >50 | 35 (7.5) |
| I don't know | 36 (7.7) |
| Did not answer | 1 (0.2) |
| **Underlying conditions <sup>c</sup>** |  |
| Yes | 154 (24.7) |
| No | 464 (74.4) |
| Did not answer | 6 (0.9) |
| **Have you already received one of the COVID-19 vaccines** |  |
| Yes | 401(64.3) |
| No | 223 (35.7) |
| Did not answer | 0 (0) |
| **Brand of vaccine received <sup>d</sup>** |  |
| Covishield | 365 (91) |
| Covaxin | 34 (8.5) |
| Did not answer | 2 (0.5) |
| **Previous confirmed or suspected COVID diagnosis** |  |
| Yes | 134 (21.5) |
| No | 439 (70.3) |
| I don't know | 0 (0) |
| Did not answer | 51 (8.2) |

<sup>a</sup> Health-facility setting occupations that are (1) patient-facing: nurses, medical doctors, clinic workers, hospital paramedical workers and (2) non-patient facing: admission/reception, housekeeping/cleaning staff, laboratory personnel, non-healthcare frontline workers. Non-health-facility setting occupations are those where respondents worked in public health capacities outside of health-facility settings.
<sup>b</sup> Among respondents who reported having an occupation in a health-facility setting (**n = 469**)
<sup>c</sup> Underlying conditions among respondents reporting “Yes” include one or more of the following: asthma, cardiovascular disease, chronic lung disease, chronic renal disease, diabetes mellitus, and hypertension (**n = 154**)
<sup>d</sup> Among respondents who reported having received at least the first dose vaccine at the time of survey completion (**n = 401**)

Among respondents, 95.0% felt confident regarding protecting themselves from SARS-CoV-2 infection, including access to recommended personal protective equipment (PPE) (87.5%) and updated information related to COVID-19 (86.4%). Reliability on government sources, healthcare providers or scientific journal articles for COVID-19 related information was reported by 66.8%, while confidence in social media for this information was reported by only 4.5%.

### COVID-19 Vaccine acceptance

Among all respondents included in the analysis, 84.1% (525/624) were accepting of the COVID-19 vaccines currently approved by the national government. An approved COVID-19 vaccine was received by 64.3% (401/624) at the time of administration of the survey. Amongst the 223 respondents who had not yet received a COVID-19 vaccine, 63.2% (141/223) were willing to accept an approved COVID-19 vaccine. Amongst these 223 respondents, those who were unwilling to accept a vaccine were younger (median age, 32 vs. 36 years, p<0.05), more often had a previously confirmed or suspected COVID-19 diagnosis (25.7% vs 19.2%, p=0.28), were less likely to have underlying conditions (20.7% vs 25.2%, p=0.45), and fewer perceived a high degree of susceptibility to COVID-19 (19.5 vs 29.1%, p=0.08) compared to those who were willing to accept a COVID-19 vaccine (Supplementary table 1).

### Factors influencing the decision to receive a COVID-19 vaccine

Eighty-three percent (518/624) of respondents believed that a vaccine would help control the spread of COVID-19. Further, 89.7% (560/624) felt that it would still be important to receive a COVID-19 vaccine even though cases were declining in India at the time of administration of the survey, prior to the second surge. Amongst respondents who had not yet received a COVID-19 vaccine at the time of survey administration, 50% (95/190) suggested that they would prefer to wait for a few months before taking a novel vaccine, 50% (111/223) suggested that their decision to accept the vaccine would be influenced by the prevalence of infection at the time of vaccination, and 71% (159/223) suggested that their decision would be influenced by the side effects of the vaccine. After adjusting for differences in participant characteristics, using multivariable logistic regression analyses, the factors independently associated with an increased likelihood of accepting the vaccine were, advancing age (aOR 3.50 [95% CI, 1.04-11.76] for those aged above 45 years), availability of evidence that the vaccine was effective (aOR 3.78 95% CI 1.15-12.38]), and provision of a vaccine free of cost (aOR 2.63 [95% CI, 1.06-6.50]). Other demographic factors such as age, gender, occupation (health-facility vs. non-health-facility setting), type of institution where they worked (public vs. private), presence of underlying comorbidities and perceived degree of susceptibility to COVID-19 were not associated with vaccine acceptance (Table 2). Further, the respondents’ decisions to accept a COVID-19 vaccine were not significantly associated with factors such as recommendations by the ministries of health or global organizations, vaccine-related discussions on media and social media, their peers’ acceptance of the vaccine, and reported vaccine side effects (Table 2).

**Table 2.**
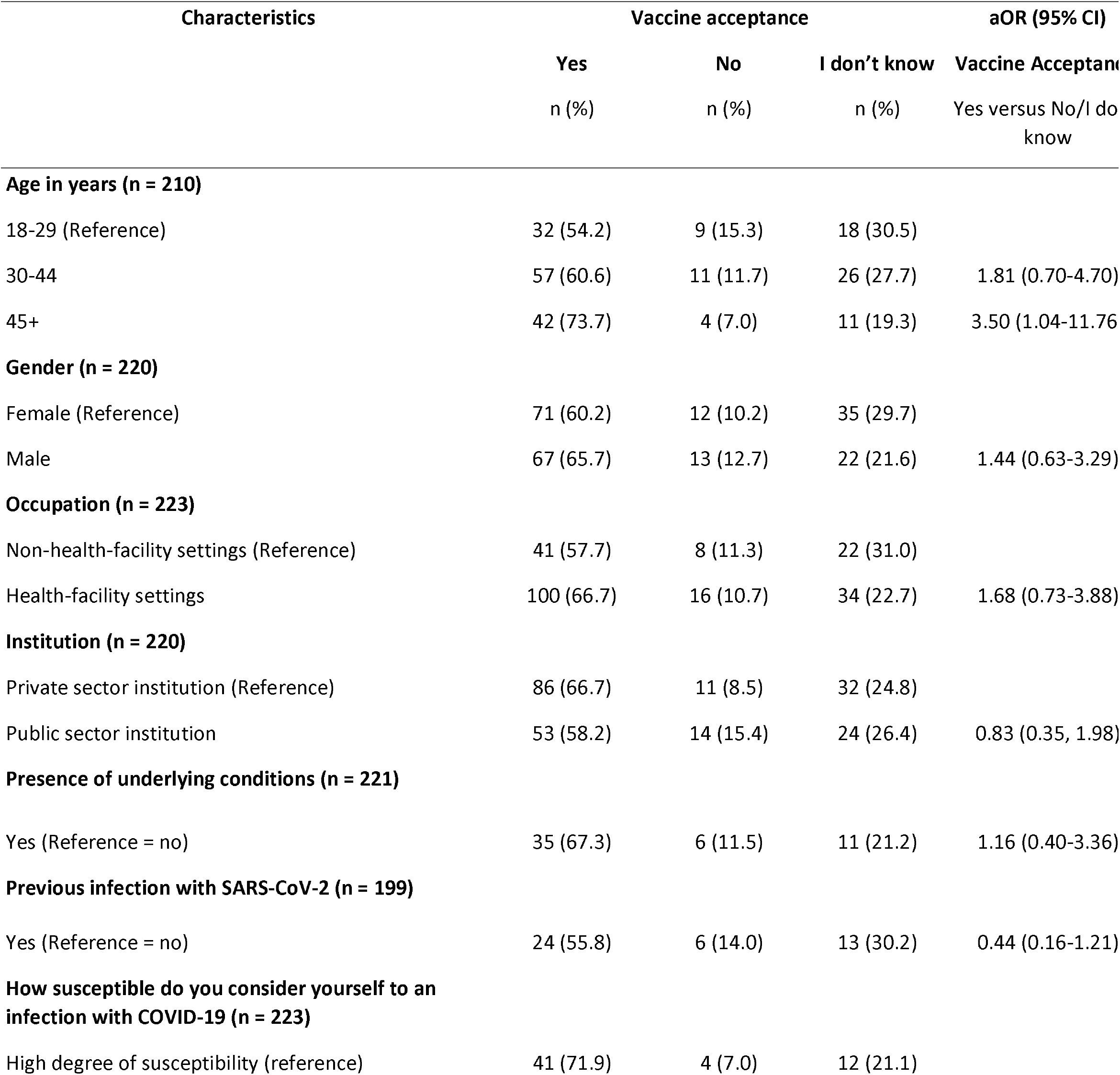
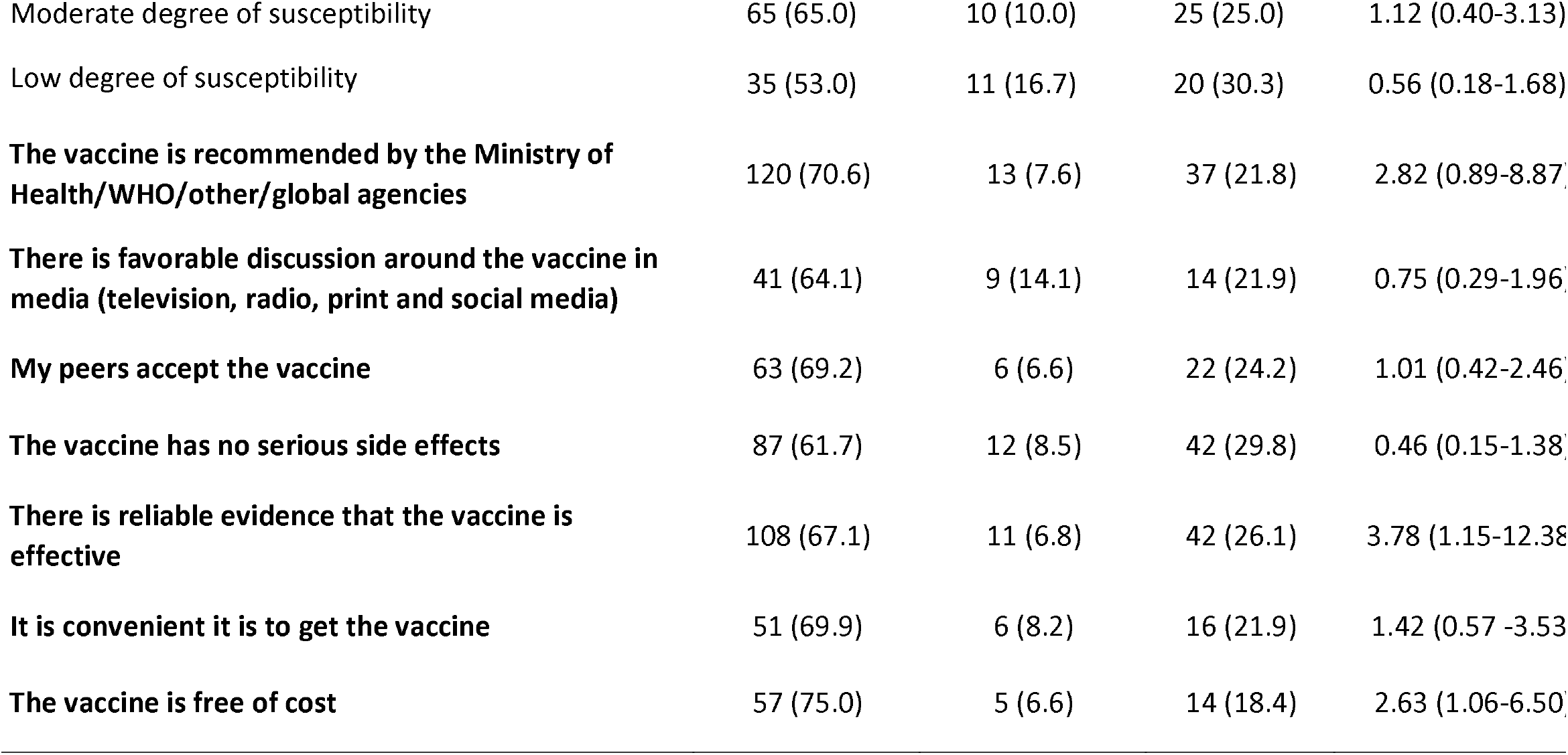
Factors influencing the decision to accept a COVID-19 vaccine currently approved by the government among respondents who had not yet received a vaccine at the time of survey administration **(n = 223)**

### Perceptions surrounding vaccine preparedness in healthcare facilities

Overall, most respondents were confident about healthcare systems being equipped to efficiently rollout COVID-19 vaccines to the general population. Confidence in different domains was reported by respondents (sufficient number of trained staff: 81.0%; procuring adequate supplies: 77.0%; efficient storage and deployment: 79.6%).

## Discussion

In a representative sample of HCWs across India, our study conducted a week into the launch of India’s COVID-19 vaccination campaign found that overall vaccine acceptance was high at 84%, but amongst those who had not yet received the vaccine at the time of survey administration, vaccine acceptance was 63%. Most respondents felt adequately empowered to find appropriate information about the disease and vaccine, indicated having knowledge on infection prevention, and believed that a vaccine would help in curtailing the spread of the infection. HCWs were more likely to accept the vaccine if they were older, had access to reliable evidence of vaccine effectiveness, and were provided with vaccine that was free of cost.

Being arbiters of scientific and public health information, HCWs are critical in promoting vaccine acceptance. As vaccine recipients themselves, and vaccine advocates, they can play a pivotal role in helping with vaccine messaging, addressing vaccine hesitancy among the public, and promoting rapid vaccine coverage within countries and globally. Vaccinating HCWs is also particularly important, as they are at increased risk of infection acquisition due to the nature of their work. In a recent systematic review on COVID-19 vaccine hesitancy worldwide, which included a total of 31 studies, eight studies were pertinent to HCWs; it was found that vaccine acceptance rates ranged from 27.7% in the Democratic Republic of the Congo to 78.1% in Israel (6). A nationwide survey of undergraduate medical students in India conducted at the launch of the vaccination campaign in India found that 89.2% participants were willing to accept a vaccine (13). In our study, we found that amongst the HCWs who had not yet received a COVID-19 vaccine, acceptance was at 63%. Studies of vaccine acceptance amongst HCWs have shown that factors associated with acceptance include male sex (14, 15), older age (15-17), education level (16, 17), perceiving a high risk for infection (14), friends/peers/relatives having had previous COVID-19 infection (14, 18), trust with their government’s measures (14) and occupational exposure (15). Recent data have also suggested that individuals were influenced by the effectiveness of COVID-19 vaccines, and they would be less willing to accept vaccines with low efficacy (19, 20).

Cost considerations were important as was found in our study. Affordability has often been cited as a barrier to vaccine uptake (21). India’s income inequality ranks among the highest in the world and it is largely due to regional disparities, rich-poor divide and the urban-rural gap (22). As the federal government in India has decided to make COVID-19 vaccines available free of cost to all adults beginning June 21, 2021, it is possible that this will continue to motivate individuals to accept the COVID-19 vaccines, and improve the equity in accessing vaccines (23). Following this change, data indicate that the average number of daily doses administered have exceeded those documented in earlier periods, and these are encouraging signs (24).

Additionally, our survey demonstrated that nearly 50% of respondents’ decisions to accept a COVID-19 vaccine would be impacted by the prevalence of COVID-19 in India at the time they were offered a vaccine. India’s recent surge of cases and deaths during the second wave of COVID-19 cases, partially attributed to the increase in viral variants (25), highlights the need for vaccination programs to outpace the development of viral variants. Scaling up vaccine manufacturing and rolling out vaccines as quickly and widely as possible are critical to protect citizens before they are exposed to the virus and new variants (26). Achieving a high vaccine coverage level to reach herd immunity is also critical in combatting the delta variant, which is on the rise and is the most infectious variant known so far (27), and largely responsible for the second surge of COVID-19 cases in India, as well as outbreaks across the world (28).

While identifying factors associated with acceptance is critical for public health messaging, identifying factors that contribute to hesitancy is similarly critical to ending the pandemic. Although HCWs in the United States have acknowledged the importance of vaccination to public health, many still express apprehensions over serious adverse effects from the COVID-19 vaccine (29). In our study, a majority of respondents shared that their decision to accept a vaccine would depend on whether the vaccine does not have serious side effects; however, this did not achieve statistical significance when analyzed in the subgroup of HCWs who had not yet received the vaccine. Skepticism over the fast-tracked vaccine development and approval process has also contributed to hesitancy amongst HCWs worldwide (15, 30, 31). In our study, 50% of HCWs stated that their decision to accept a vaccine would be influenced by whether the vaccine has been in use for over six months.

It is critical to effectively design communication strategies to address the unique concerns HCWs and general populations may have about the COVID-19 vaccine. Vaccinated HCWs have a noticeable positive impact on individuals’ decisions to accept a COVID-19 vaccine (32); therefore, messaging that emphasizes that majority of vaccination experiences are positive, and highlights acceptance among HCWs, will lead to more community support of COVID-19 vaccines. Communication campaigns need to leverage strategies to design community-informed social and behavioral messages, target positive factors associated with acceptance (33), and address factors that impact hesitancy. Efforts that put people at the center of efforts may culminate in increased vaccine uptake (34).

Accessible and equitable global roll-out of vaccines for COVID-19 is essential to curb the spread of the pandemic, protect against severe disease, and drive herd immunity. According to the World Health Organization’s COVID-19 vaccine introduction toolkit (35), successful deployment of the vaccines will involve regulatory preparedness, identifying target populations and delivery strategies, handling supply and related logistics, adequate human resources and training and providing vaccine specific resources. Most respondents in our study felt that health facilities in India were adequately prepared to undertake the massive endeavor of the largest vaccination drive ever attempted by any country. Since the launch of the COVID-19 vaccination campaign in India on January 16, 2021, 17.5% of the population have received at least one dose, and 3.7% are fully vaccinated as of June 24,2021 (36, 37). It will remain critical to sustain and scale up these efforts as the vaccination campaign advances in order to successfully control the spread of the pandemic.

Our study has some limitations that are worth noting. The survey was initiated one week after the initial rollout of the COVID-19 vaccines in India when information, options, and perceptions were rapidly changing. We acknowledge the possibility of selection bias; those who were respondents to our survey may have had strong opinions, whether positive or negative, about vaccination. Responses by the HCWs may have been affected by social desirability bias. Further, this was a cross-sectional survey administered at a single time-point, with no longitudinal follow-up; willingness to get vaccinated may change over time and be influenced by factors such as newer knowledge and the emergence of variants. This survey was conducted during a time when COVID-19 cases were on the decline in India, prior to the second surge, which could have impacted perceptions of individuals responding to our survey. Despite these limitations, our cross-sectional study is strengthened by the fact that we received responses from a diverse group of HCWs from across India.

In conclusion, in this cross-sectional survey conducted at the onset of the COVID-19 vaccination rollout in India, we found that amongst a representative group of HCWs in India, overall attitudes towards COVID-19 vaccination and preparedness were positive, with the majority willing to accept a vaccine. Favorable vaccine efficacy and provision of vaccination free of cost were significantly associated with acceptance. As HCWs serve as ambassadors for evidence-based medical interventions, and they are critical in promoting vaccine acceptance amongst the general population, it is essential to design targeted interventions to improve vaccine acceptance amongst this population. Successful rollout of COVID-19 vaccines in an effort to mitigate the effects of the pandemic will require leveraging public health allies to improve trust and confidence in communities across the country—an endeavor that needs to start with healthcare workers.

## Data Availability

Data are submitted along with the manuscript

## Tables

**Supplementary table 1.** Comparison of HCWs by willingness to accept a COVID-19 vaccine, amongst those who had not received a vaccine at the time of survey administration (n=223)

| Characteristics | Total<br>(n=223) | Yes (willing to<br>accept a vaccine)<br>n = 141 | No/I don't<br>know (unwilling<br>to accept a<br>vaccine)<br>n = 82 | p-value |
| --- | --- | --- | --- | --- |
| **Median age (IQR) (years)** | 35 (29-46) | 36 (30-49) | 32 (27-42) | 0.02 |
| **Gender** |  |  |  |  |
| Male | 102 (46.4) | 67 (48.6) | 35 (42.7) | 0.39 |
| Female | 118 (53.6) | 71 (51.4) | 47 (57.3) |  |
| **Type of institution** |  |  |  |  |
| Private sector | 129 (58.6) | 86 (61.9) | 43 (53.1) | 0.20 |
| Public sector | 91 (41.4) | 53 (38.1) | 38 (46.9) |  |
| **Occupation <sup>a</sup>** |  |  |  |  |
| Health-facility setting | 150 (67.9) | 100 (70.9) | 50 (62.5) | 0.19 |
| Non-health-facility setting | 71 (32.1) | 41 (29.1) | 30 (37.5) |  |
| **Presence of underlying**<br>**conditions <sup>b</sup>** | 52 (23.3) | 35 (25.2) | 17 (20.7) | 0.45 |
| **Previous confirmed or**<br>**suspected COVID-19**<br>**diagnosis** | 43 (19.2) | 24 (19.2) | 19 (25.7) | 0.28 |
| **How susceptible do you consider yourself to an infection with COVID-19** |  |  |  |  |
| High degree of susceptibility | 57 (25.6) | 41 (29.1) | 16 (19.5) | 0.08 |
| Moderate degree of<br>susceptibility | 100 (44.8) | 65 (46.1) | 35 (42.7) |  |
| Low degree of susceptibility | 66 (29.6) | 35 (24.8) | 31 (37.8) |  |

<sup>a</sup> Health-facility setting occupations that are (1) patient-facing: nurses, medical doctors, clinic workers, hospital paramedical workers and (2) non-patient facing: admission/reception, housekeeping/cleaning staff, laboratory personnel, non-healthcare frontline workers. Non-health-facility setting occupations are those where respondents worked in public health capacities outside of health-facility settings.
<sup>b</sup> Underlying conditions among respondents reporting “Yes” include one or more of the following: asthma, cardiovascular disease, chronic lung disease, chronic renal disease, diabetes mellitus, and hypertension.

## Acknowledgements

The authors acknowledge support from the Indian Association of Preventive and Social Medicine (IAPSM).

